# Correcting Algorithmic Bias in Machine Learning Prediction of Healthcare utilization in India

**DOI:** 10.1101/2025.09.07.25335256

**Authors:** John Tayu Lee, Vincent Cheng-Sheng Li, Sheng Hui Hsu, Tzu-Pin Lu, Charlotte Wang, Arokiasamy Perianayagam, Kanya Anindya, Rifat Atun

## Abstract

**Objective:** This study investigates how historical disparities in healthcare access influence machine learning (ML) predictions of healthcare utilization among older adults in India through algorithmic bias. We examine the extent to which standard ML models underestimate utilization in disadvantaged populations and quantify the resulting distortion in national-level cost projections.

**Methods:** Using data from 55,698 respondents in the Longitudinal Ageing Study in India (LASI), we trained two sets of ML models to predict outpatient and inpatient utilization: one on the full population (Model 1), and another on a subsample with met healthcare needs (Model 2). Gradient Boosting was selected as the best-performing algorithm. To interpret model predictions and identify key drivers of healthcare utilization, we applied SHapley Additive exPlanations (SHAP). We compared model outputs across socioeconomic subgroups and extrapolated predicted utilization to national population estimates using WHO-CHOICE unit costs.

**Findings:** Model 1 consistently underestimated healthcare utilization relative to Model 2, particularly among lower-income and caste-identified groups. Overall, outpatient and inpatient predictions from Model 2 were 8.92 (95% CI: 8.87–8.99)% and 9.59 (9.28–9.85)% higher, respectively. Nationally, this translated to an underestimation of I$390.7 (391.2–391.5) million in outpatient care and I$88.4 (86.2–90.1) million in inpatient care. The largest gaps were concentrated in the poorest and most marginalized subgroups. The SHAP analysis suggests that self-rated health (SRH), economic status (MPCE), and chronic conditions are consistently influential in predicting outpatient and inpatient visits, with some shifts in feature importance between models.

**Conclusion:** Machine learning models trained on unadjusted population data lead to algorithmic bias and risk perpetuating structural inequities by underrepresenting unmet need. Models based on fulfilled care scenarios yield more equitable and accurate projections.

## INTRODUCTION

The rise of machine learning in analysis of public health data has opened new frontiers for predictive modeling and policy design ^1^. However, these models are only as reliable as the data they are built upon. Health data often carry embedded historical inequities that reflect systemic barriers in access and utilization, particularly among the poor ^2^. As a result, models trained on such data produce algorithmic bias (defined as “the instances when the application of an algorithm compounds existing inequities in socioeconomic status, race, ethnic background, religion, gender, disability or sexual orientation to amplify them and adversely impact inequities in health systems”) ^3^ may perpetuate, or even amplify, longstanding disparities in healthcare delivery and resource allocation ^4^.

Take the task of predicting healthcare utilization as an example. When training data underrepresent the healthcare needs of socioeconomically disadvantaged groups due to past access limitations, algorithmic bias occurs and machine learning models are likely to learn skewed patterns ^5^. These models risk systematically underestimating the true care requirements of the lower socioeconomic population groups—even in hypothetical settings where barriers are removed—thus compounding disadvantage through under-prediction ^6^.

A prominent example of algorithmic bias is illustrated by Obermeyer et al. (2019), who found that a widely used commercial algorithm predicted future healthcare costs rather than actual health status or illness severity ^4^. Because Black patients, on average, incur lower healthcare costs than White patients at equivalent levels of illness—due to structural disparities in access and treatment—the algorithm significantly underestimated their health needs ^5^. As a result, fewer Black patients were identified for high-risk care programs, despite having greater underlying medical burden.

Algorithmic bias produced due to under-representativeness of historical data in machine learning applications for healthcare and public health refers to the embedded prejudices present in datasets used to train AI models. These biases often stem from systemic inequities in the delivery of healthcare services, demographic imbalances, and historical contexts that have contributed to disparities in care. For example, datasets may capture a legacy of unequal treatment toward racial and ethnic minority populations, reinforcing the AI models leading to these existing inequities ^7,8^. Such historical bias might exist in several forms, including the underrepresentation of specific demographic groups in training datasets, which can cause AI models to underperform for them. Furthermore, missing data and misclassification errors may distort the model accuracy, potentially worsen existing disparities in health outcomes ^5,9^.

To address this issue, this study investigates the manifestation and impact of historical biases that produce algorithmic bias in predictive models of healthcare utilization using data from the Longitudinal Ageing Study in India (LASI). We analyze how disparities in healthcare access for lower-income groups shape the observed data and, consequently, the predictions made by standard algorithms. Going beyond the identification of bias, we quantify its effect by comparing predictions derived from a full population sample versus a subgroup with fulfilled healthcare needs.

Specifically, we performed the following analyses: (1) we compared machine learning predictions of healthcare utilization using population-wide data versus data from individuals with fully met healthcare needs. (2) we assessed the extent to which standard models underestimate utilization among socioeconomically disadvantaged groups. (3) we quantified the implications of such bias in national-level healthcare cost estimations.

## RESULTS

### Descriptive Statistics

The study analyzed data from 55,698 individuals aged 45 years and older, sampled from the nationally representative Longitudinal Ageing Study in India (LASI). The results are presented in table 1. Within the sample, 18% of respondents identified as Scheduled Caste, 17% as Scheduled Tribe, 38% as Other Backward Class, and 28% as Other or No Caste. To assess economic differences, participants were categorized into five equally sized groups based on Monthly Per Capita Expenditure (MPCE).

**Table 1.** Summary of Descriptive Statistics (Full Data)

|  | **Respondent**<br>**No. (%)** | **Average Frequency of**<br>**Outpatient Care** | **Average Frequency of**<br>**Inpatient Care** |
| --- | --- | --- | --- |
| **Overall** | **55,698** | **2.4651** | **0.0844** |
| **Caste** |  |  |  |
| Scheduled Caste | 9,819 (18%) | 1.1361 | 0.0697 |
| Scheduled Tribe | 9,404 (17%) | 2.9011 | 0.0854 |
| Other Backward Class | 21,103 (38%) | 2.6365 | 0.0890 |
| Other/No Caste | 15,372 (28%) | 2.8119 | 0.0870 |
| **MPCE** |  |  |  |
| Lowest | 11,286 (20%) | 1.7831 | 0.0495 |
| Lower Middle | 11,319 (20%) | 2.2366 | 0.0636 |
| Middle | 11,137 (20%) | 2.5493 | 0.0839 |
| Upper Middle | 11,038 (20%) | 2.8376 | 0.0959 |
| Highest | 10,918 (20%) | 2.9445 | 0.1311 |

To examine the pattern of healthcare utilization, we first calculated and compare its average per group. The results in Table 1 show that patterns of healthcare utilization exhibited significant disparities across both caste and income lines. Among the full sample, respondents reported an average of 2.47 outpatient visits and 0.08 inpatient episodes in the previous year. Scheduled Tribe participants reported the highest average of outpatient care (2.90 visits), while Scheduled Caste respondents had the lowest (1.14 visits). Similarly, a socioeconomic gradient was observed in inpatient care: individuals in the lowest MPCE quintile reported an average of 0.050 annual admissions, compared to 0.131 in the highest quintile.

Table 2 shows that among those whose healthcare needs were met—derived by excluding those with unmet need from the full dataset (n=4421)—the average utilization levels were consistently higher. In this subgroup, outpatient care rose to 2.68 visits per year and inpatient episodes increased to 0.09. Notably, while access barriers were minimized in this group, social disparities in utilization persisted, with higher rates of service use still concentrated among Scheduled Tribe and high-income individuals.

**Table 2.** Summary of Descriptive Statistics (Subset Data)

|  | **Respondent**<br>**No. (%)** | **Average Frequency of**<br>**Outpatient Care** | **Average Frequency of**<br>**Inpatient Care** |
| --- | --- | --- | --- |
| **Overall** | **51,277** | **2.6776** | **0.0917** |
| **Caste** |  |  |  |
| Scheduled Caste | 8,437 (16%) | 1.3222 | 0.0811 |
| Scheduled Tribe | 8,723 (17%) | 3.1276 | 0.0921 |
| Other Backward Class | 19,704 (38%) | 2.8237 | 0.0954 |
| Other/No Caste | 14,413 (28%) | 2.9990 | 0.0928 |
| **MPCE** |  |  |  |
| Lowest | 9,932 (19%) | 2.0262 | 0.0563 |
| Lower Middle | 10,332 (20%) | 2.4503 | 0.0697 |
| Middle | 10,321 (20%) | 2.7509 | 0.0905 |
| Upper Middle | 10,383 (20%) | 3.0166 | 0.1020 |
| Highest | 10,309 (20%) | 3.1184 | 0.1388 |

### Disparities in Unmet Health Needs Among Socioeconomic Groups

We then move on to perform. The results of statistical tests on the unmet healthcare need differences are presented in Table 3 which shows that the share of older adults with an unmet healthcare need varies significantly across both caste and MPCE groups. Scheduled Caste respondents exhibit the highest unmet-need prevalence (14.3 %), more than double that of the other/no caste (6.2 %), while the lowest MPCE quintile (11.9 %) far exceeds the highest quintile (5.6 %). Large chi-square statistics (𝒳^2^ = 622.5 for caste; 413.5 for MPCE, *p* < 0.001) confirm that these differences are unlikely due to chance, although the corresponding Cramér’s V values (0.106 and 0.086) indicate the associations are small in magnitude.

**Table 3.** Chi-square & Cramér’s V for Unmet-Need Proportions by Caste and MPCE

|  | Unmet Need No. (% within group) | Chi-square | p-value | Cramér's V |
| --- | --- | --- | --- | --- |
| **Caste** |  | 622.5 | <0.001 | 0.106 |
| Scheduled Caste | 1,382 (14.3%) |  |  |  |
| Scheduled Tribe | 681 (7.2%) |  |  |  |
| Other Backward Class | 1,399 (6.6%) |  |  |  |
| Other/No Caste | 959 (6.2%) |  |  |  |
| **MPCE** |  | 413.5 | <0.001 | 0.086 |
| Lowest | 1,354 (11.9%) |  |  |  |
| Lower Middle | 987 (8.7%) |  |  |  |
| Middle | 816 (7.3%) |  |  |  |
| Upper Middle | 655 (5.9%) |  |  |  |
| Highest | 609 (5.6%) |  |  |  |

### Model Performance

To evaluate predictive accuracy, seven machine learning algorithms were trained on the full sample (Model 1), and the results are presented in Table 4. For both outpatient and inpatient outcomes, Gradient Boosting clearly outperformed all other methods. It attains the lowest error—RMSE = 3.4232 visits (95% CI 3.2954–3.5548) for outpatient care, and RMSE = 0.4187 admissions (95% CI 0.3415–0.5104) for inpatient care. For outpatient visits, Random Forest is the next-best method (RMSE = 3.4444 (95% CI 3.3118–3.5689)), whereas the linear and Lasso regressions, together with the two neural nets (DNN, FCN), cluster around RMSE ≈ 3.5, and SVR performs worst (RMSE = 3.5857 (95% CI 3.4416–3.7286)).

**Table 4.**
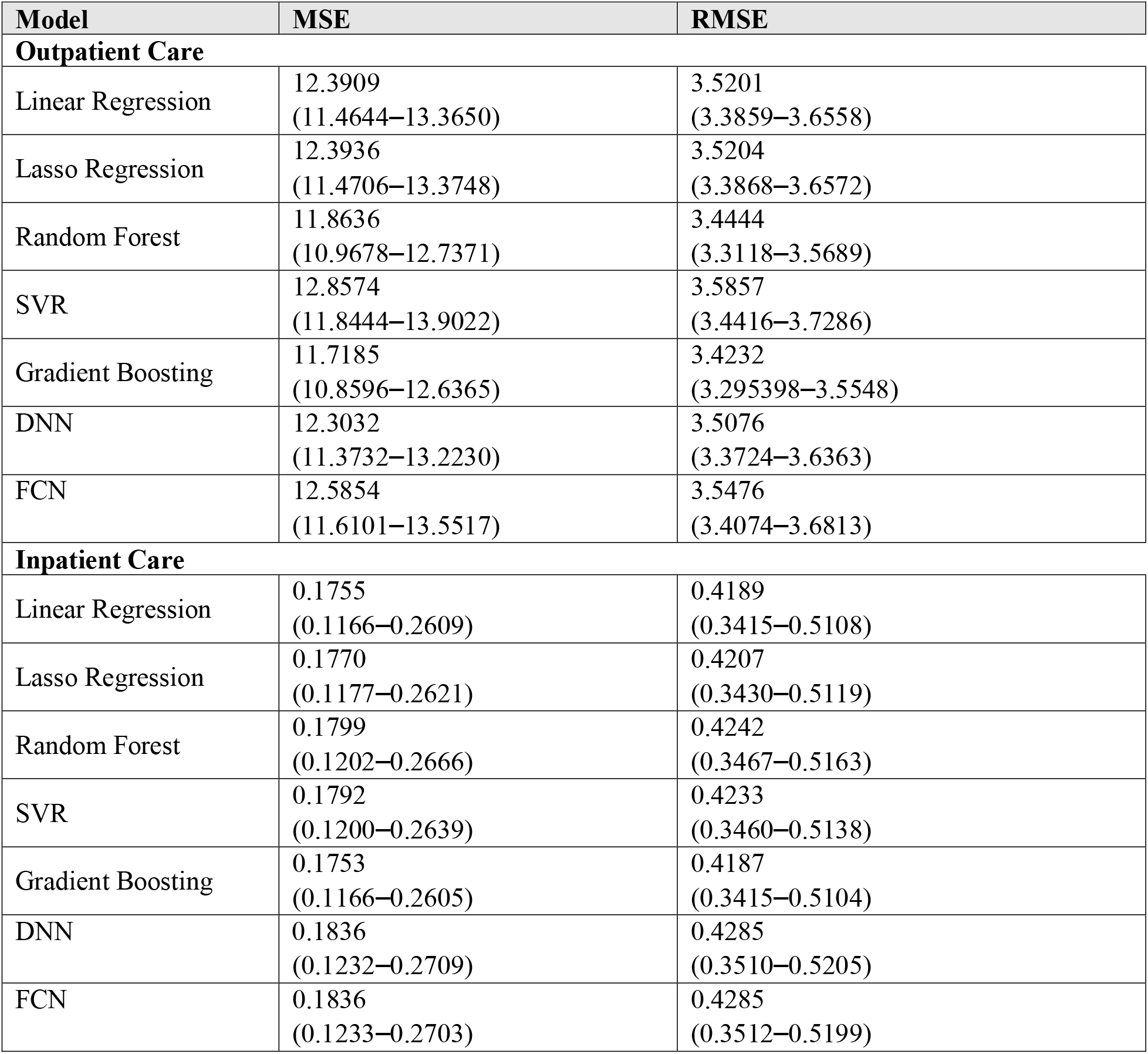
Model Performance Metrics

For inpatient admissions the ordering changes: Gradient Boosting retains a marginal lead, but the Linear and LASSO regressions achieve comparable accuracy, whereas Random Forest, the neural networks and SVR fall to the lower tier, with RMSE ≈ 0.42. Taken together, the results indicate that ensemble tree boosting provides the most favorable bias–variance balance for both the common outpatient outcome and the infrequent inpatient outcome; consequently, Gradient Boosting was selected for all subsequent utilization analyses.

Underestimation of Utilization

Subsequent analysis in Table 5 compared the predictions of the two models with observed care use. At the population level, Model 1 predicted 137,488 (136,608-138,383) outpatient visits in the last 12 months—nearly identical to the observed total of 137,301. In contrast, Model 2 projected 149,746 (148,892–150,655) visits, reflecting a 8.92% (8.87–8.99) difference. This suggests that Model 1, trained on the full sample individuals including those with unmet needs, systematically underestimated actual demand. For inpatient care, Model 1 estimated 4,690 (4,655-4,727) admissions compared to an observed total of 4,703, while Model 2 predicted 5,140 (5,114–5,166)—a 9.59% (9.28–9.85) difference over Model 1.

**Table 5.** Group Comparison Using the Best Performing Model (Gradient Boosting)

| Group | Observed Count | Predicted Count (Model 1) | Predicted Count (Model 2) | Difference (%) |
| --- | --- | --- | --- | --- |
| **Outpatient Care** |  |  |  |  |
| Overall | 137,301 | 137,488<br>(136,608–138,383) | 149,746<br>(148,892–150,655) | 8.92<br>(8.87–8.99) |
| High MPCE | 91,861 | 91,098<br>(90,385–91,810) | 97,670<br>(96,937–98,365) | 7.21<br>(7.14–7.25) |
| Low MPCE | 45,440 | 46,390<br>(45,891–46,857) | 51,682<br>(51,186–52,227) | 11.41<br>(11.46–11.54) |
| Castes | 94,076 | 93,084<br>(92,397–93,805) | 102,470<br>(101,675–103,239) | 10.08<br>(10.04–10.06) |
| Non-Castes | 43,225 | 44,404<br>(43,969–44,861) | 47,127<br>(46,708–47,616) | 6.13<br>(6.14–6.23) |
| **Inpatient Care** |  |  |  |  |
| Overall | 4,703 | 4,690<br>(4,655–4,727) | 5,140<br>(5,114–5,166) | 9.59<br>(9.28–9.85) |
| High MPCE | 3,424 | 3,270<br>(3,241–3,302) | 3,308<br>(3,288–3,328) | 1.16<br>(0.79–1.48) |
| Low MPCE | 1,279 | 1,420<br>(1,401–1,437) | 1,820<br>(1,807–1,833) | 28.20<br>(27.58–28.95) |
| Castes | 3,366 | 3,244<br>(3,213–3,273) | 3,628<br>(3,607–3,647) | 11.81<br>(11.43–12.23) |
| Non-Castes | 1,337 | 1,445<br>(1,425–1,467) | 1,510<br>(1,496–1,524) | 4.45<br>(3.85–4.98) |

Furthermore, these discrepancies were more pronounced among disadvantaged groups. For outpatient care, the lowest MPCE quintile saw a 11.41% (11.46–11.54) increase in predicted visits from Model 2 compared to Model 1, while the highest quintile had a 7.21% (7.14–7.25) increase. Similarly, among caste-identified groups, Model 2 estimated 10.08% (10.04–10.06) more outpatient visits than Model 1, compared to a 6.13% (6.14–6.23) difference in non-caste groups. In inpatient care, Model 2 predictions were 28.20% (27.58–28.95) higher than Model 1’s for the lowest MPCE quintile, and 11.81% (11.43–12.23) higher among caste-identified groups. These differences reveal a clear pattern: models that do not account for access disparities tend to underpredict utilization most significantly in marginalized populations.

Figure 1 extends the findings in Table 5 by depicting mean predicted utilization for each socioeconomic stratum. Relative to Model 1, Model 2 produces uniformly higher estimates— roughly 0.2 additional outpatient visits per person (a 7–11% increase, peaking in the low-MPCE group) and about 0.02 additional inpatient admissions (up to a 30% relative rise for the same group). The virtually imperceptible confidence bars indicate that these increases are systematic rather than sampling variation and are most pronounced among economically disadvantaged households.

**Figure 1.**
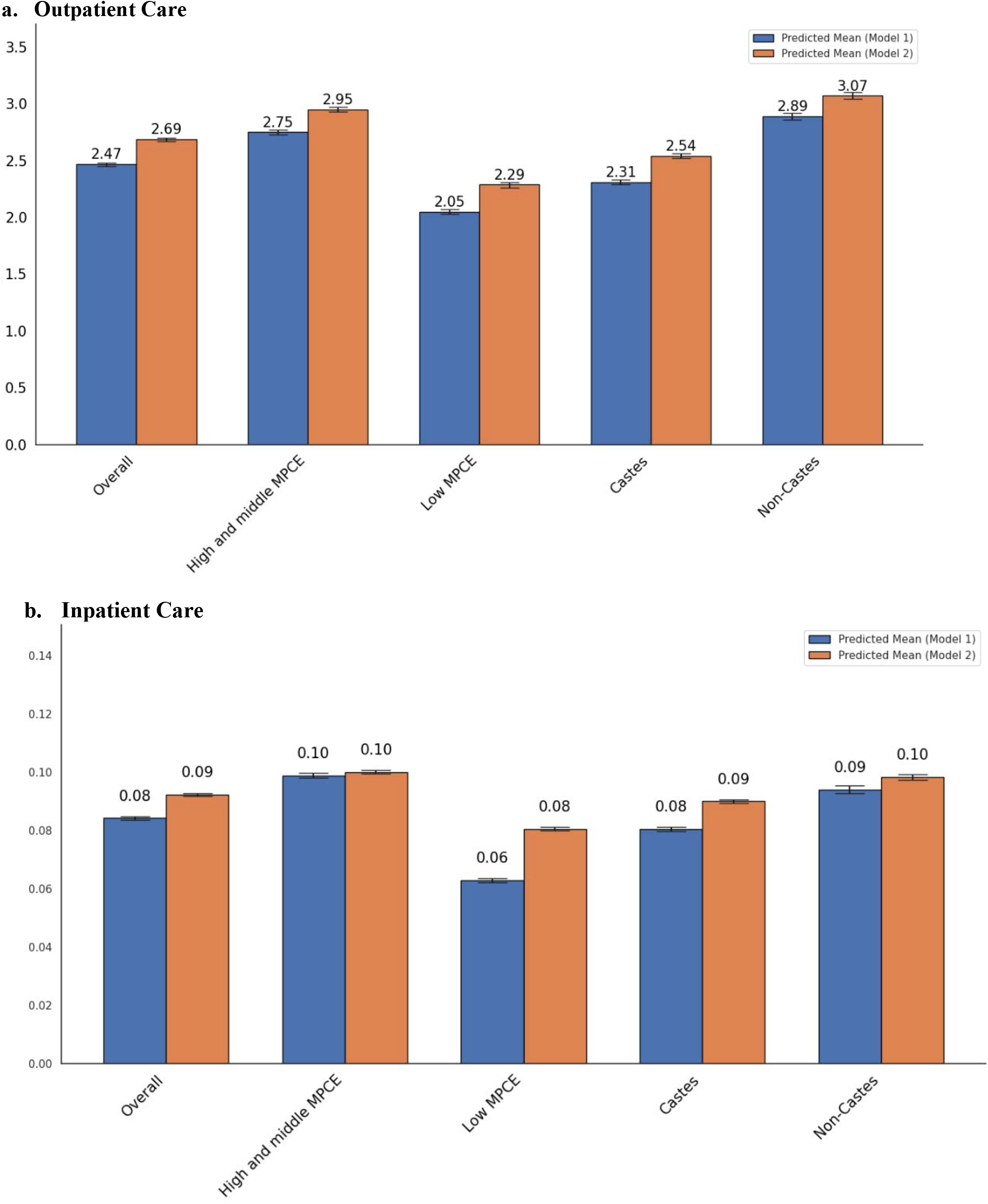
Expected Value Analysis

### Cost Implications for India’s Health System

To evaluate the financial impact of these biases, we applied the WHO-CHOICE unit costs for outpatient (I$5.58) and inpatient (I$34.41) services to each model’s predictions and extrapolated them to India’s population aged 45 and above in 2017 (approximately 318 million people).

In Table 6, for outpatient care, both observed data and Model 1 predicted a total cost of approximately I$4.38 (4.35-4.41) billion. Model 2, however, estimated a cost of I$4.77 (4.75– 4.80) billion, reflecting a national-level underestimation of I$390.7 (95% CI 391.2–391.5) million. Cost underestimations were concentrated among vulnerable groups, including I$168.7 (95% CI 168.8–171.2) million in the lowest MPCE quintile and I$299.2 (295% CI 95.7–300.7) million among caste-identified populations.

**Table 6.** National-Level Outpatient Care Cost Estimates by Population Subgroup in India

**a. Outpatient Care**
| Group | Estimated Population Aged 45+ | Observed Cost (I\$) | Predicted Cost (Model 1, I\$) | Predicted Cost (Model 2, I\$) | Difference (I\$) |
| --- | --- | --- | --- | --- | --- |
| Overall | 318,168,118 | 4,376,479,934 | 4,382,428,599<br>(4,354,393,716–4,410,971,376) | 4,773,177,696<br>(4,745,942,113–4,802,149,740) | 390,749,097<br>(391,178,364–391,548,397) |
| High MPCE | 189,039,778 | 2,928,076,616 | 2,903,745,558<br>(2,881,028,531–2,926,452,710) | 3,113,234,405<br>(3,089,871,561–3,135,381,534) | 209,488,848<br>(208,843,029–208,928,824) |
| Low MPCE | 129,128,340 | 1,448,403,591 | 1,478,683,042<br>(1,462,768,471–1,493,558,834) | 1,647,353,241<br>(1,631,551,608–1,664,724,675) | 168,670,200<br>(168,783,137–171,165,841) |
| Castes | 230,357,419 | 2,998,679,882 | 2,967,053,276<br>(2,945,172,546–2,990,046,838) | 3,266,244,324<br>(3,240,893,631–3,290,739,321) | 299,191,048<br>(295,721,084–300,692,483) |
| Non-Castes | 87,810,699 | 1,377,800,356 | 1,415,375,323<br>(1,401,503,475–1,429,938,056) | 1,502,188,759<br>(1,488,818,378–1,517,749,915) | 86,813,436<br>(87,314,904–87,811,859) |

**b. Inpatient Care**
| Group | Estimated Population Aged 45+ | Observed Cost (I\$) | Predicted Cost (Model 1, I\$) | Predicted Cost (Model 2, I\$) | Difference (I\$) |
| --- | --- | --- | --- | --- | --- |
| Overall | 318,168,118 | 924,435,721 | 921,812,116<br>(915,026,791–929,226,842) | 1,010,240,139<br>(1,005,166,016–1,015,413,462) | 88,428,023<br>(86,186,620–90,139,225) |
| High MPCE | 189,039,778 | 673,031,650 | 642,758,014<br>(636,966,210–649,046,383) | 650,213,130<br>(646,386,218–654,190,236) | 7,455,116<br>(5,143,853–9,420,008) |
| Low MPCE | 129,128,340 | 251,404,051 | 279,054,102<br>(275,432,968–282,477,439) | 357,757,453<br>(355,163,003–360,372,597) | 78,703,351<br>(77,895,158–79,730,035) |
| Castes | 230,357,419 | 661,630,982 | 637,716,230<br>(631,647,956–643,326,666) | 713,048,970<br>(708,903,442–716,850,261) | 75,332,740<br>(73,523,595–77,255,486) |
| Non-Castes | 87,810,699 | 262,804,704 | 284,095,886<br>(280,129,968–288,391,349) | 296,731,287<br>(294,079,912–299,507,564) | 16,191,772<br>(11,116,215–13,949,944) |

Inpatient care followed a similar pattern. While Model 1 estimated total national inpatient costs at I$921.8 (915.0–929.2) million—closely aligned with observed costs of I$924.4 million— Model 2 estimated I$1010.2 (1005.2–1015.4) million, a difference of I$88.4 (86.2–90.1) million. Disaggregated estimates showed the greatest underestimation in the lowest MPCE group (I$78.7, 95% CI 77.9–79.7) million) and among caste groups (I$75.3, 95% CI 73.5–77.3 million).

### Feature Importance Using SHAP

Figure 2 displays layered-violin SHAP summaries for Model 1 and 2. The top panels refer to outpatient-care predictions, the bottom to inpatient care. Along each horizontal axis, positive SHAP values raise the predicted number of visits, negatives lower it; the violin width shows how often a given impact occurs, and the blue-to-red colors map low-to-high feature values.

**Figure 2.**
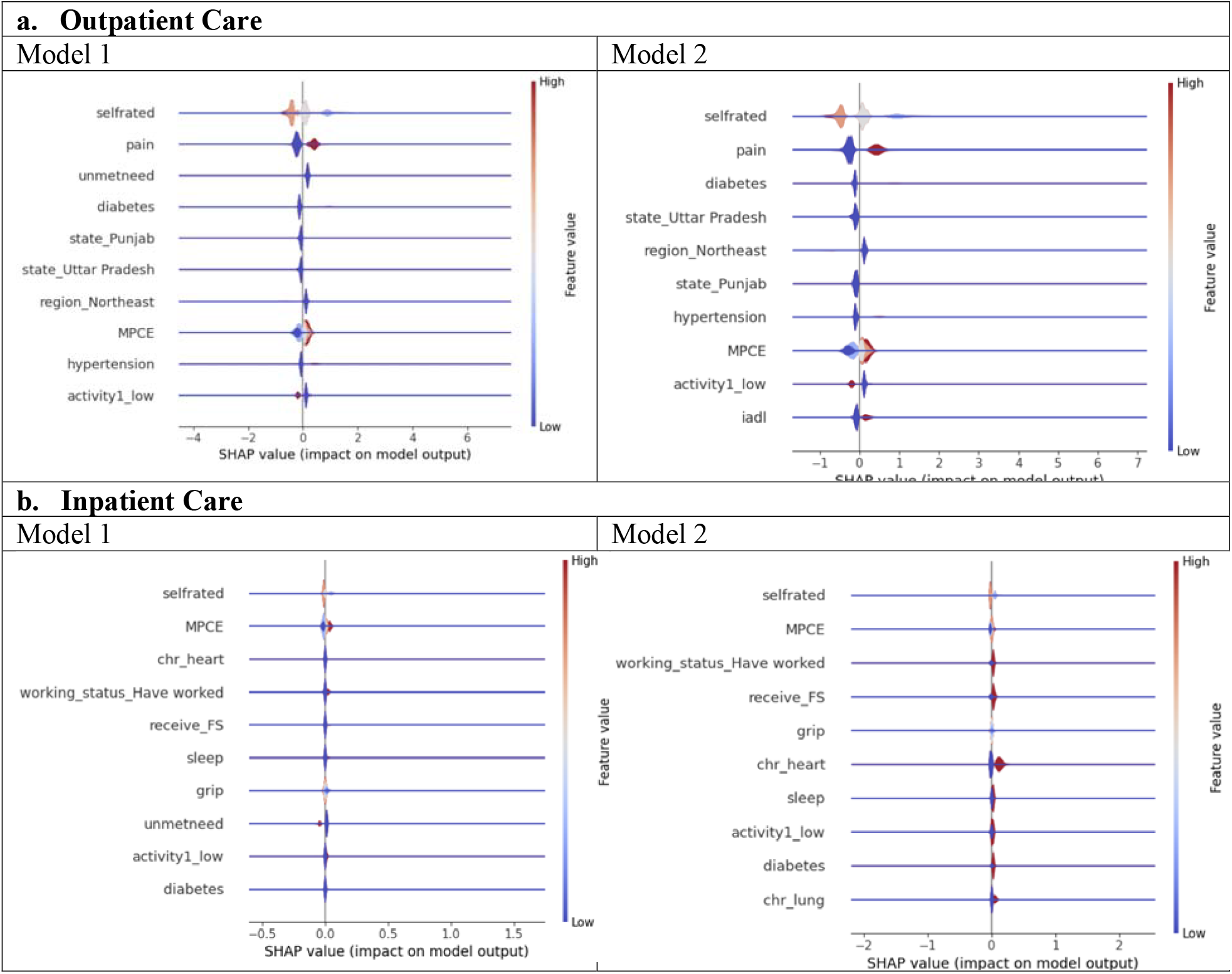
SHAP Value Analysis

For outpatient care (upper panels), the rank order of determinants is largely consistent across the two specifications, though with notable shifts. Adverse self-rated health (SRH) exerts the greatest positive influence on predicted visit counts, followed by pain and diabetes. In Model 1, the indicator for unmet need emerges as a prominent predictor; as anticipated, this term is absent in Model 2 once observations with unmet need are excluded, and hypertension shows slightly higher contributor. The geographic pattern also shifts: in Model 1, residence in Punjab and Uttar Pradesh is associated with higher utilization, whereas after the sample restriction the strongest regional effect originates from the North-East, followed by Uttar Pradesh. In both models, higher economic status (high MPCE) consistently increase the predicted number of outpatient visits.

For inpatient care (bottom panels), SRH remains the most influential predictor, followed by MPCE and a history of chronic heart disease. In Model 1 the indicator for unmet need and a prior employment history both contribute substantial positive effects; in Model 2, the unmet-need term is absent by design and a chronic-lung condition enters the top-ten list, whereas the impacts of employment status and financial support (receive FS) persist. In every panel the majority of SHAP values cluster near zero—reflecting baseline predictions that amount to only fractions of an admission—so the long, slender tails of the violins are generated exclusively by observations with extreme feature profiles.

To account for the complex survey design, we incorporated individual sampling weights in the sensitivity analysis. The results were consistent with those of the main analysis, suggesting the robustness of our findings (see Appendix Tables 1–4 and Figure 1).

## DISCUSSION

These findings highlight a critical methodological concern in the application of machine learning to health services research. When model are trained on population-wide data that includes historically unmet needs leads to algorithmic bias that tends to systematically underestimate healthcare utilization and associated costs, particularly for disadvantaged socioeconomic groups. By contrast, models trained on individuals with fulfilled healthcare needs offer more accurate and equitable predictions. These results call attention to the need for more nuanced model development strategies that explicitly account for disparities in healthcare access. Without such adjustments, predictive models may perpetuate or even exacerbate existing inequities by informing decisions that misallocate resources away from the populations that need them most.

Bias in training data significantly affects machine learning (ML) predictions of healthcare use or access, particularly impacting disadvantaged or underserved populations ^13^. This bias can arise from several sources, including historical inequities, incomplete data, and misclassification errors ^3,5^. Firstly, ML models trained on biased data can perpetuate and even exacerbate existing healthcare disparities ^4^. For instance, models may underdiagnose conditions in underserved populations due to historical underrepresentation or misclassification in the training data. This can lead to delayed or inadequate care for these groups ^14^. Secondly, the performance of ML models often varies across different sociodemographic groups. Studies have shown that models may have higher error rates for individuals from lower socioeconomic statuses or racial minorities, leading to unequal healthcare outcomes ^15^. This differential performance is often due to incomplete or less accurate data for these populations, which can skew predictions and resource allocation ^16^.

Our study highlights sample representativeness as a critical source of bias in applying machine learning to health system planning. When training data reflect historical inequalities or discrimination in health service use pattern, these disparities can be perpetuated in future predictions ^17^.

To mitigate these biases, several strategies have been proposed. These include preprocessing techniques like resampling and reweighting data to ensure more balanced representation, as well as post-hoc methods to adjust model outputs for fairness ^18^. Additionally, frameworks like BE-FAIR emphasize the importance of incorporating equity considerations throughout the model development and deployment process to enhance fairness and reduce disparities ^19^.

The findings underscore the critical need for caution and methodological rigor in the application of ML models for healthcare financing planning, particularly in diverse and potentially data-scarce settings like India and to prioritize Data Representativeness. The study’s focus on sample representativeness adjustment highlights a fundamental challenge in leveraging historical healthcare utilization data for future projections. Policymakers and model developers must ensure that the data used to train ML models accurately reflects the underlying population and their diverse healthcare needs ^2,4^. This includes investing in robust data collection strategies that minimize sampling bias and capture the experiences of underserved populations with unmet needs ^15^. Failure to address data representativeness can lead to biased predictions, potentially exacerbating existing health inequities in resource allocation ^13^.

When building ML models for resource allocation or financing of healthcare interventions, it is crucial to explicitly define and account for ‘desired behaviors’ related to healthcare utilization. Historical data may reflect existing barriers to access or underutilization due to various socio-economic factors. Simply predicting future utilization based on such biased data risks perpetuating these undesirable patterns ^17^. Policy interventions aimed at promoting equitable access and encouraging appropriate healthcare seeking behaviors should be integrated into the modeling process, potentially through techniques that weight or adjust data to reflect these desired outcomes ^18,20^.

Our research suggests that bias mitigation strategies should begin with a thorough examination of data sampling issues. Addressing potential biases at the data acquisition and preprocessing stages is likely more effective than solely relying on post-modeling bias correction techniques ^19^. Policymakers should encourage and invest in research that develops and evaluates methods for identifying and mitigating sampling bias in healthcare data ^15^. This includes promoting the use of diverse data sources and the development of techniques to re-weight or augment underrepresented groups in the training data ^16^.

Given the potential for ML models to influence healthcare resource allocation, transparency and explainability are paramount. Policymakers should advocate for the development of models that are interpretable, allowing stakeholders to understand the factors driving predictions and identify potential sources of bias ^21^. This fosters trust and facilitates informed decision-making regarding the deployment of ML in healthcare financing ^22^.

The implementation of ML-driven healthcare financing strategies should be accompanied by continuous monitoring and evaluation. This includes tracking the accuracy and fairness of the models over time, as well as assessing their impact on health equity and access ^23^. Feedback loops and mechanisms for model recalibration based on real-world outcomes are essential to ensure that these tools contribute to a more just and efficient healthcare system ^2^.

The study underscores the fairness of ML must be embedded at the core of healthcare financing strategies. Bias in predicting healthcare-related outcomes often originates from unrepresentative training data shaped by historical disparities particularly in caste and socioeconomic groups ^24^. To address this issue, policymakers should prioritize representative sampling and enforce rigorous subgroups testing to reveal and correct bias, especially for the marginalized ^25^. Our study suggests that ML model trained on unmet-need-inclusive data significantly underpredict healthcare utilization up to 28.20 (27.58–28.95) % for the poorest. This underestimation could distort the allocation of resources or perpetuate inequity. Therefore, data correction through representative sampling, equity weighting, and the creation of benchmark models based on fulfilled healthcare needs should become standard policy practice^26^. Further, the applications of ML in population health must be evaluated for fairness across subgroups, with transparency in bias metrics and ongoing model recalibration, enabling ML to evolve from being just a predictive tool into a mechanism that actively mitigates structural inequities ^27^. The reliability of ML applications in policy making hinges not only on algorithmic accuracy but on the social and statistical integrity of its data foundations ^28^.

In conclusion, ML creates a major potential for optimizing healthcare financing, but its application requires a cautious and nuanced approach. Policymakers must prioritize data quality and representativeness, explicitly consider desired healthcare behaviors, and recognize data sampling as a critical point for bias mitigation ^1^. The reliability of ML model outputs rests not on algorithms alone, but on the integrity of the data that are used to develop them. Without this foundation, predictive precision risks masking structural inequities rather than addressing them ^13,17^.

## METHODS

### Study Design and Objectives

This study quantifies prediction bias in healthcare utilization models by comparing predictions based on the full sample of older Indian adults with those derived from a subsample of individuals whose healthcare needs were met. By isolating this subgroup, we approximate a less biased representation of true healthcare needs, allowing a more accurate assessment of underestimation caused by historical disparities in healthcare access.

### Data Source

We conducted a cross-sectional analysis using the 2017–2018 wave of the Longitudinal Ageing Study in India (LASI), a nationally representative survey of individuals aged 45 and older across all Indian states and union territories. LASI captures a broad range of information on health, social, and economic well-being, enabling robust aging and public health analyses. From the initial 72,250 individuals surveyed, we excluded 6,350 due to proxy responses or age below 45. Following data cleaning to remove incomplete records, the final analytical sample comprised 55,698 participants.

### Identification of the Met-Need Subgroup

To identify individuals whose healthcare needs were fulfilled, we constructed a binary indicator of unmet need. Respondents who did not visit health care facility and did not consult to any health care provider in the past year for reasons other than “did not get sick” were classified as having unmet healthcare needs. Please refer to Appendix Table for definitions of all variables used in this study. Those who did not report any unmet need are considered to have had their needs met. This met-need subgroup was used to train an alternative predictive model intended to reflect care-seeking behaviors in the absence of access barriers. We compared socioeconomic and caste group differences in prevalence of unmet needs using Chi-squared test (Table 3).

### Outcome Variables

Two primary outcomes were examined: (1) Outpatient visits: The number of times a respondent received medical care or consultation (including home visits) from a health care provider, including doctor, AYUSH practitioner, etc., over the previous 12 months; (2) Inpatient visits: The number of overnight hospital or long-term care admissions in the past 12 months.

### Predictors

Our 76 predictors span multiple domains. Demographic variables include age, gender, state-region, rural-urban residence, marital status, and living-alone status. Socio-economic factors cover caste, MPCE quintile, employment type, and pension receipt and amount. Health indicators comprise diagnoses of 20+ chronic diseases, symptom-based conditions (e.g., pain, sleep disturbance, malaria, dengue), functional limitations (ADL/IADL difficulties, low vision, grip strength, denture use, assistive aids), BMI category, central obesity, low cognition, depression and self-rated health. Behavioral risks include smoking, smokeless tobacco, drinking pattern, physical-activity level, yoga/meditation and severe food insecurity. Social context is captured by decision-making role, financial transfers given and received, grandchild care, organizational membership, discrimination experience and overall life-satisfaction. Categorical variables were one-hot encoded and continuous variables z-standardized before model training.

### Data Preprocessing

Continuous variables were standardized, and categorical variables were processed using one-hot encoding. Two analytical datasets were created: (1) full dataset: all respondents (n=55,968); (2) met-need subgroup dataset: refers to the subset of the full dataset, excluding all individuals with unmet healthcare needs (n=51,277).

### Machine Learning Models

We implemented seven machine learning algorithms to predict healthcare utilization: Linear Regression, Lasso Regression, Random Forest, Support Vector Regression (SVR), Gradient Boosting, DNN and FCN.

Two versions of each algorithms were trained: (1) Model 1: Trained on the full population dataset, (2) Model 2: Trained on the met-need subgroup dataset. Hyperparameter tuning was performed using randomized search to optimize model performance. Model performance was assessed using Mean Squared Error (MSE), Root Mean Squared Error (RMSE) and R^2^ on the test set; for every metric we attached 95 % confidence intervals obtained from 1,000 bootstrap resamples of the actual–predicted observation pairs. All analyses were conducted using Python 3.12.0.

### Subgroup and Cost Analysis

To assess potential underestimation biases, predictions from Model 1 and Model 2 were compared across socioeconomic subgroups, including caste categories and quintiles of Monthly Per Capita Expenditure (MPCE).

To estimate healthcare expenditures, we applied unit cost estimates from the WHO-CHOICE 2010 framework, expressed in 2010 international dollars (PPP I$) ^10^. Specifically, the cost per outpatient visit was set at I$5.58, and the cost per inpatient bed-day at I$34.41 ^11,12^. National-level estimates were generated by applying LASI-derived population weights to the Indian population aged 45 and above in 2017 (approx. 318 million individuals). The estimation process involved the following multi-step procedure:

#### 1. Estimating Subgroup Population Size

Let *P*_*total*_ represent the total Indian population aged ≥ 45 years in 2017. The estimated population size of subgroup *k*, denoted *P*_*k*_ was calculated using the observed subgroup proportion in the LASI dataset:

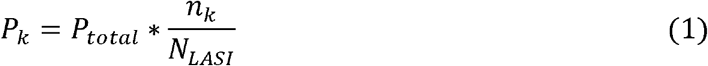

where *n*_*k*_ is the number of individuals in subgroup *k* within the LASI sample, and *N*_*LASI*_ is the total number of LASI respondents.

### 2. Average Healthcare Utilization

Mean healthcare utilization for each subgroup was computed separately for outpatient visits and inpatient bed-days. Let 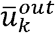 and 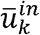 denote the average number of outpatient cares and inpatient cares per person in subgroup *k*, respectively. These values were directly estimated from individual-level data:

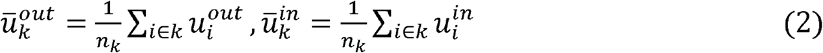

Where 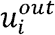 and 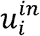 refer to the reported number of outpatient visits and inpatient bed-days, respectively, for individual i in subgroup *k*. To simplify, we assume bed-days for all inpatient visits are one.

### 3. Subgroup-Level Healthcare Expenditures

Healthcare expenditures were then estimated for each subgroup *k* by multiplying the estimated subgroup population by the average utilization and the unit cost for each type of care:

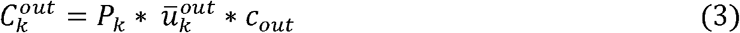

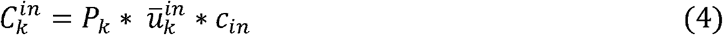

where 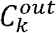 and 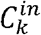 represent the total outpatient and inpatient expenditures, respectively, respectively, for subgroup *k*. The parameters 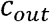 and 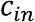 denote the unit costs per outpatient visit and per inpatient bed-day, respectively.

The total healthcare cost for subgroup *k* was then obtained by summing the outpatient and inpatient components:

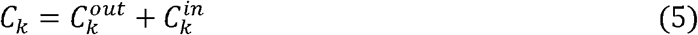

Discrepancies between the two models’ predictions were examined to assess how unmet healthcare needs influence underestimation of care utilization and costs, particularly among vulnerable and socioeconomically disadvantaged groups.

### Sensitivity Analysis

To assess the robustness of our findings and account for the complex survey design, we conducted a sensitivity analysis incorporating individual sampling weights provided by the LASI dataset. These weights help ensure that the estimates better reflect the national population structure. We retrained all models and repeated the analyses using these weights.

### Ethics Considerations

This study utilized publicly available, de-identified data from the Longitudinal Ageing Study in India (LASI), a nationally representative longitudinal survey examining the health, economic, and social well-being of India’s population aged 45 and above. The research received ethical approval from the NTU Ethical Review Board (NTU REC-No.: 202407HS010).

## Data Availability

All data produced are available online at: https://www.iipsindia.ac.in/content/LASI-data

## Code availability

Statistical analysis code in Python version 3.12.0 is available for download at: https://github.com/johntayuleeHEPI/Historical-Bias?tab=readme-ov-file

## Acknowledgements

We thank the Yushan Fellow Program by the Ministry of Education (MOE), Taiwan for the financial support. (MOE-112-YSFMN-0003-002-P1)

## Author Contributions

Concept and design: JTL and Vincent L.; Acquisition, statistical analysis, or interpretation of data: Jennifer H., JTL., Vincent L. ; *Drafting of the manuscript:* JTL., Jennifer H, and Vincent L.; *Critical review of the manuscript for important intellectual content:* All author.

## TABLE and FIGURES

**Appendix Table 1.**
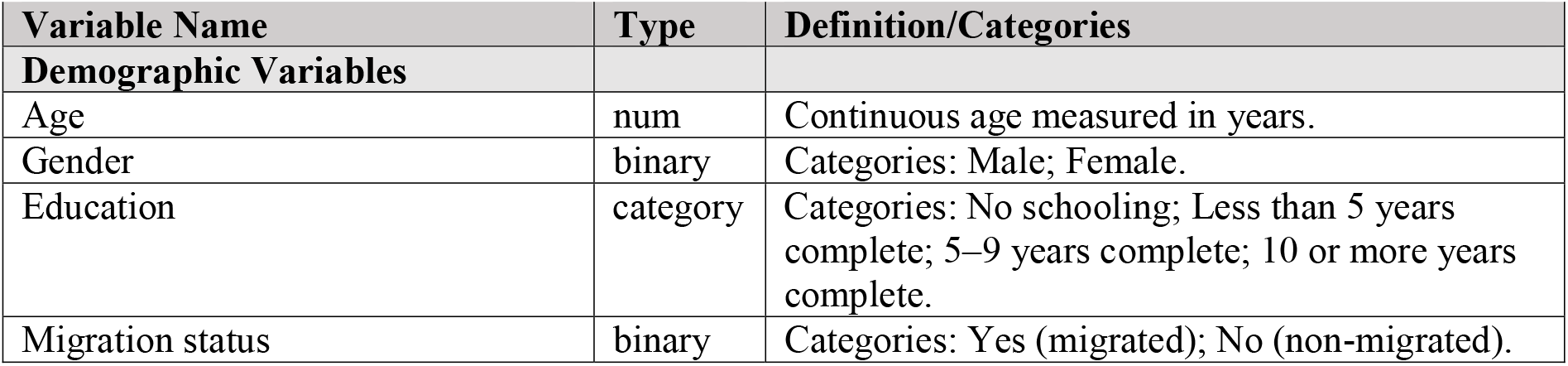
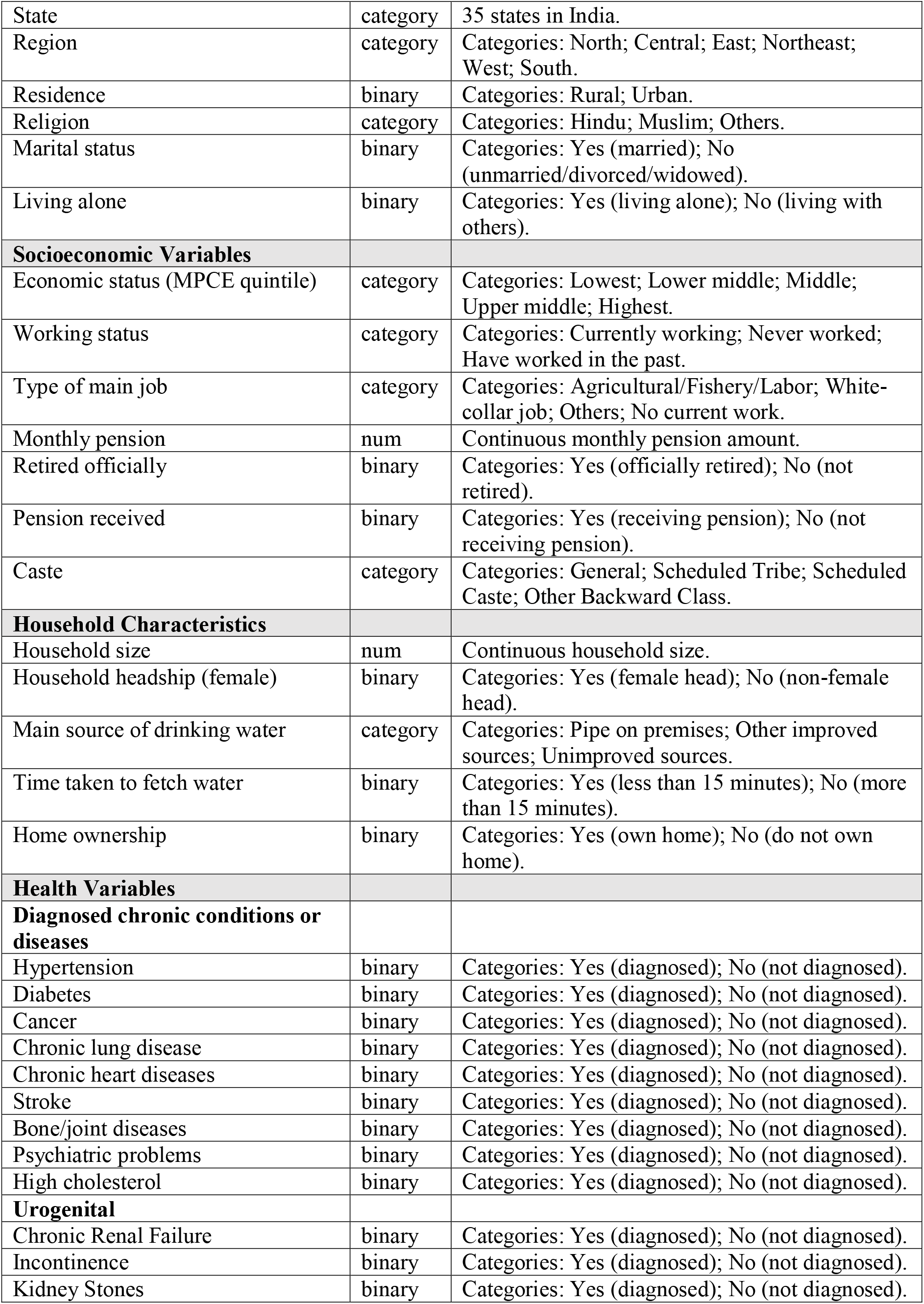
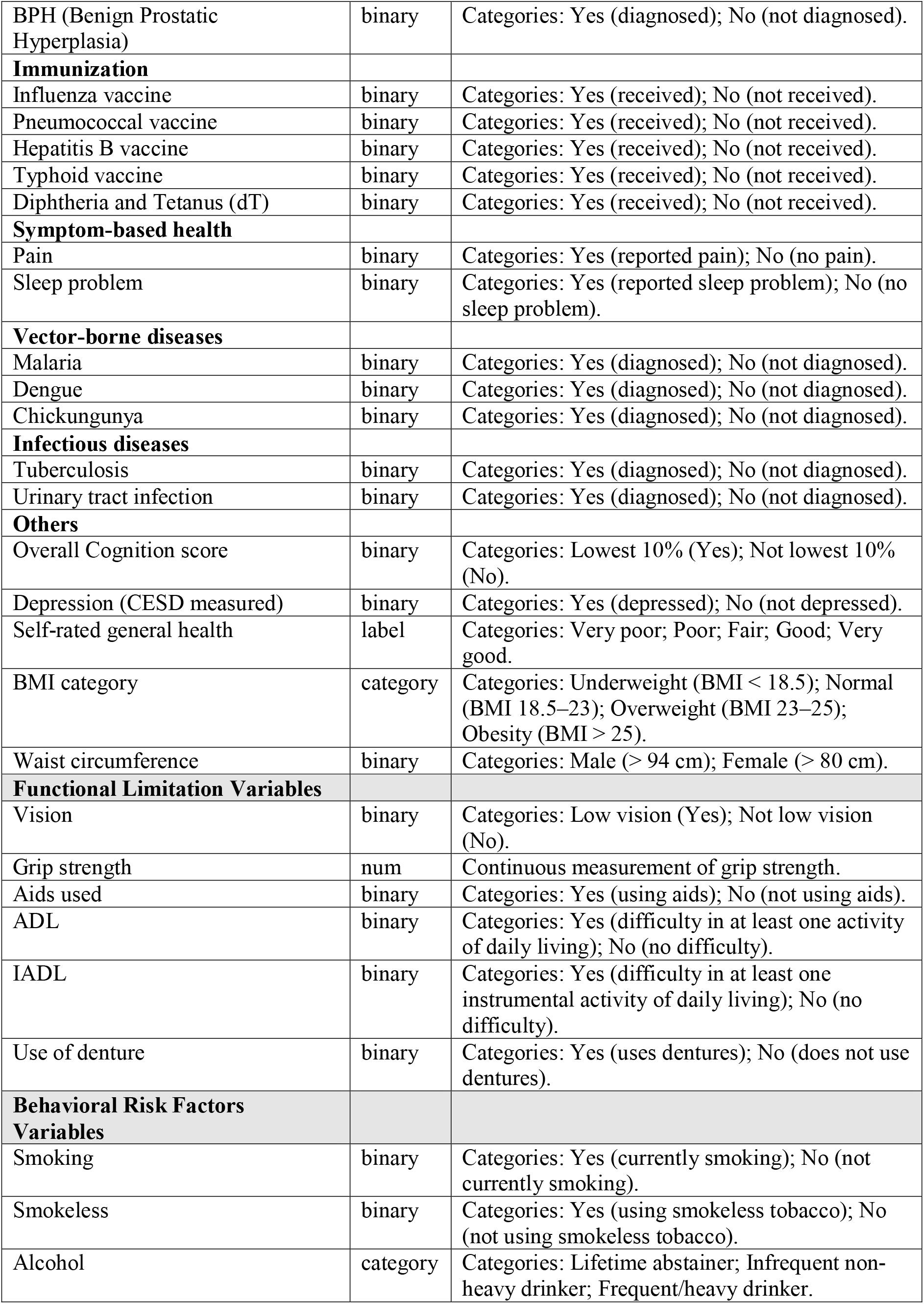
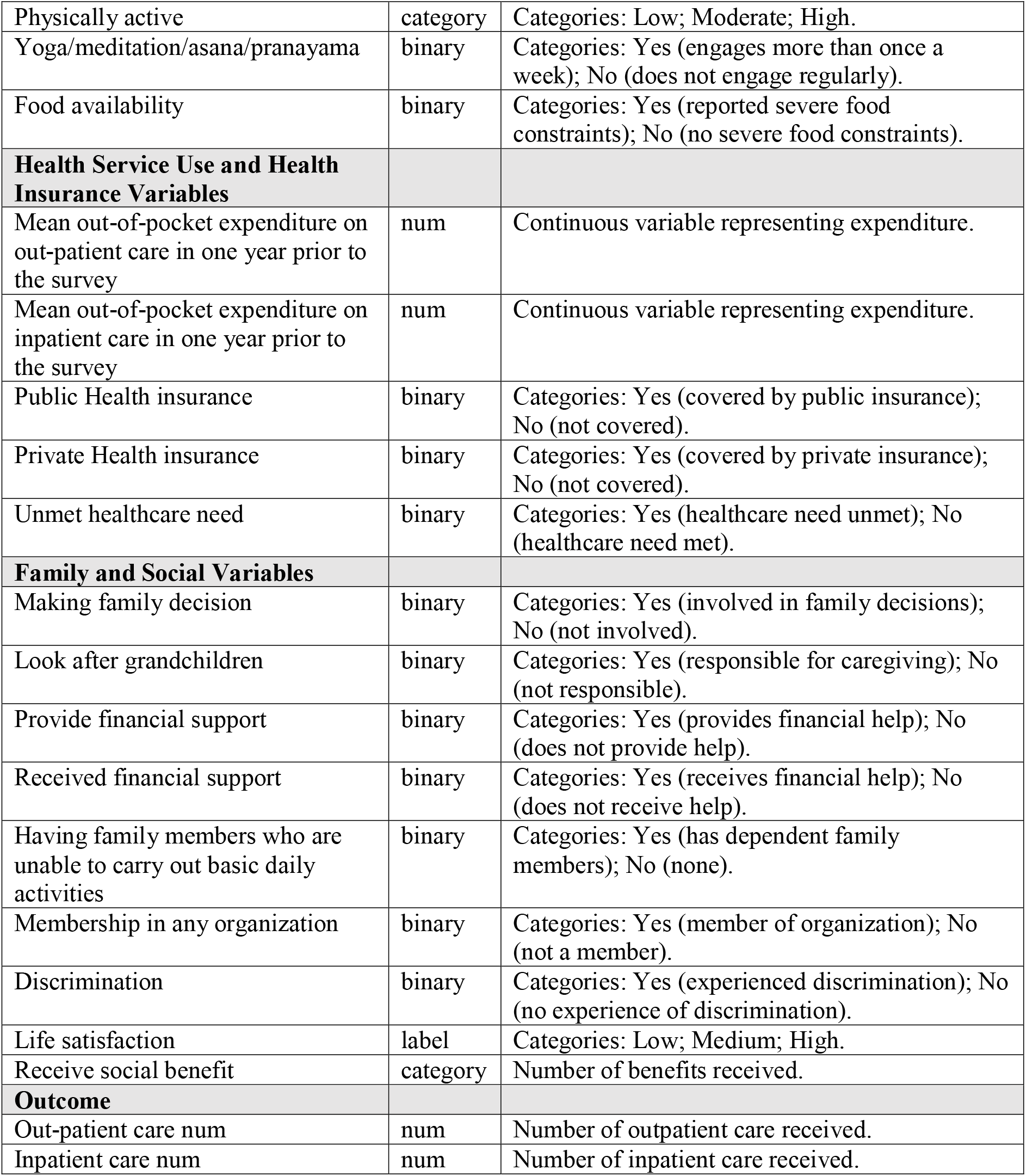
Definitions and Descriptions of Variables Used in the Analysis

**Appendix Table 2.** Weighted Model Performance Metrics

| Model | MSE | RMSE |
| --- | --- | --- |
| **Outpatient Care** |  |  |
| Linear Regression | 13.6676 (12.4136–15.0986) | 3.6970 (3.5233–3.8857) |
| Lasso Regression | 13.6811 (12.4174–15.1309) | 3.6988 (3.5238–3.8898) |
| Random Forest | 13.0854 (11.9534–14.3219) | 3.6174 (3.4574–3.7844) |
| SVR | 17.7558 (16.4210–19.2188) | 4.2138 (4.0523–4.3839) |
| Gradient Boosting | 13.3782 (12.1451–14.6905) | 3.6576 (3.4850–3.8328) |
| DNN | 14.2772 (12.8600–15.7283) | 3.7785 (3.5860–3.9659) |
| FCN | 13.6624 (12.3841–14.9963) | 3.6963 (3.5191–3.8725) |
| **Inpatient Care** |  |  |
| Linear Regression | 0.1766 (0.1201–0.2550) | 0.4203 (0.3466–0.5050) |
| Lasso Regression | 0.1771 (0.1201–0.2560) | 0.4208 (0.3465–0.5059) |
| Random Forest | 0.1881 (0.1336–0.2643) | 0.4337 (0.3655–0.5141) |
| SVR | 0.2345 (0.1767–0.3140) | 0.4842 (0.4204–0.5604) |
| Gradient Boosting | 0.2139 (0.1441–0.3034) | 0.4625 (0.3797–0.5508) |
| DNN | 0.1883 (0.1310–0.2684) | 0.4339 (0.3619–0.5181) |
| FCN | 0.1806 (0.1236–0.2611) | 0.4250 (0.3515–0.5110) |

**Appendix Table 3.** Group Comparison Using the Best Performing Weighted Model

| Group | Observed Count | Predicted Count (Model 1) | Predicted Count (Model 2) | Difference (%) |
| --- | --- | --- | --- | --- |
| **Outpatient Care (Random Forest)** |  |  |  |  |
| Overall | 137,301 | 158,088 (155,715–160,444) | 168,865 (166,331–171,282) | 6.82 (3.67–10.00) |
| High MPCE | 91,861 | 102,592 (100,302–104,759) | 107,648 (105,461–109,687) | 4.93 (0.67–9.36) |
| Low MPCE | 45,440 | 56,601 (55,461–57,765) | 61,924 (60,753–63,305) | 9.41 (5.17–14.14) |
| Castes | 94,076 | 110,449 (108,320–112,546) | 118,530 (116,264–120,655) | 7.32 (3.30–11.39) |
| Non-Castes | 43,225 | 47,921 (46,983–48,918) | 50,555 (49,601–51,578) | 5.50 (1.40–9.78) |
| **Inpatient Care (Linear Regression)** |  |  |  |  |
| Overall | 4,703 | 4,695 (4,606–4,790) | 5,163 (5,063–5,263) | 9.96 (5.70–14.27) |
| High MPCE | 3,424 | 3,471 (3,395–3,544) | 3,696 (3,624–3,779) | 6.48 (2.24–11.31) |
| Low MPCE | 1,279 | 1,214 (1,160–1,268) | 1,426 (1,368–1,487) | 17.50 (7.84–28.27) |
| Castes | 3,366 | 3,371 (3,287–3,448) | 3,666 (3,585–3,750) | 8.73 (3.98–14.11) |
| Non-Castes | 1,337 | 1,324 (1,274–1,379) | 1,496 (1,440–1,551) | 13.01 (4.46–21.71) |

**Appendix Table 4.**
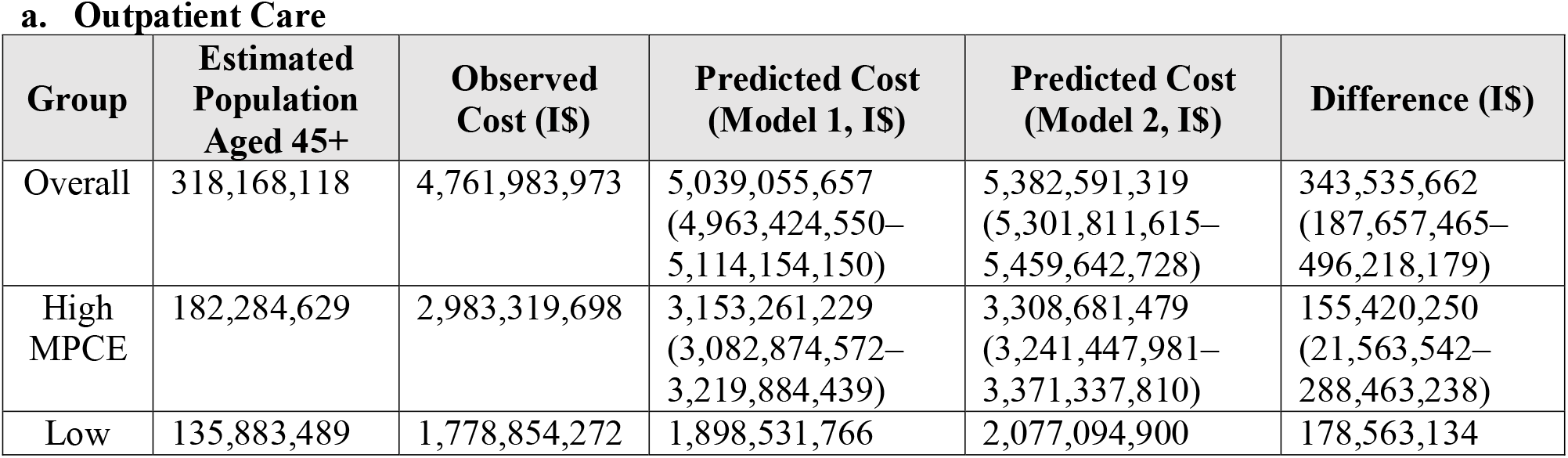
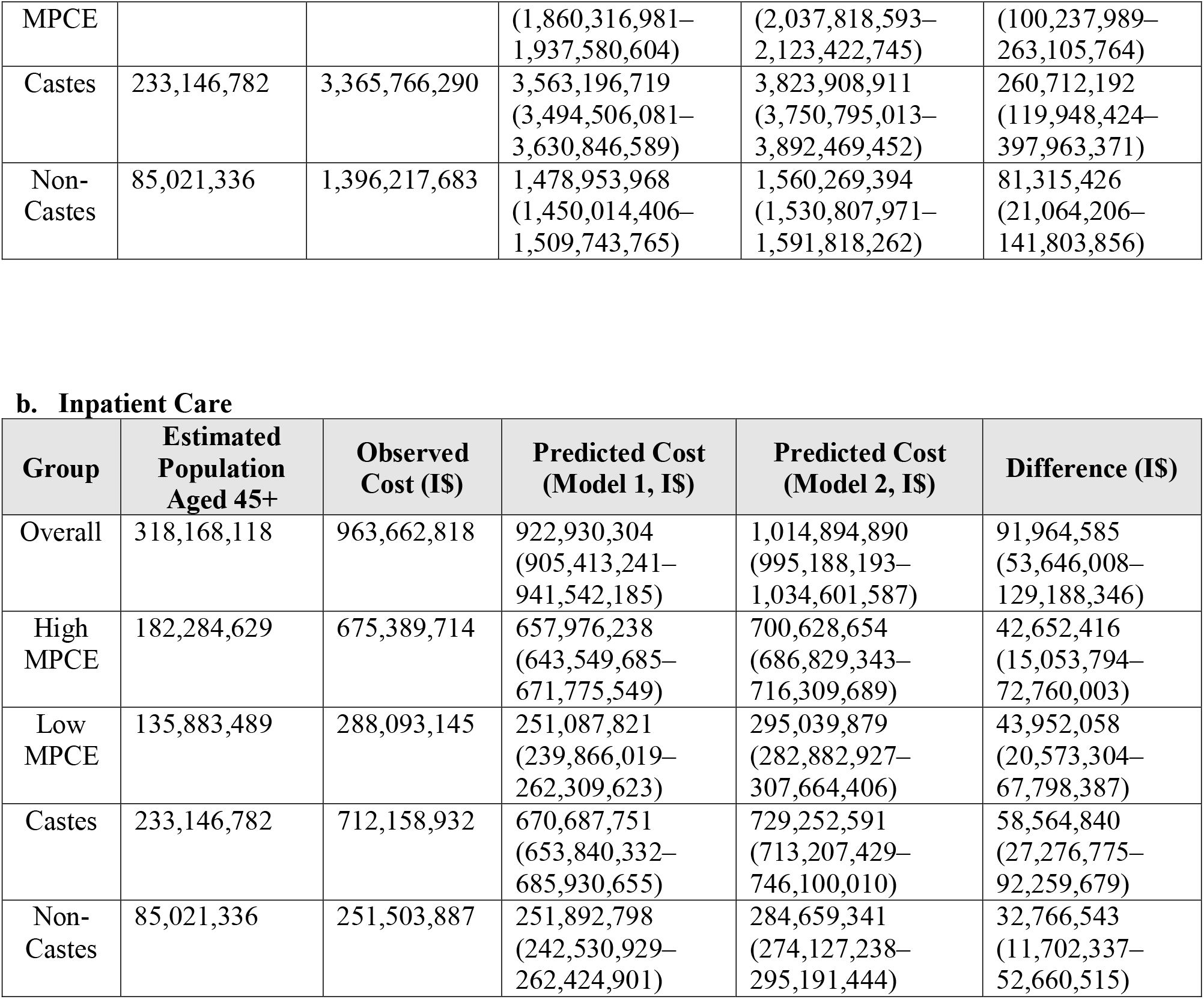
Weighted National-Level Outpatient Care Cost Estimates by Population Subgroup in India

**Appendix Figure 1.**
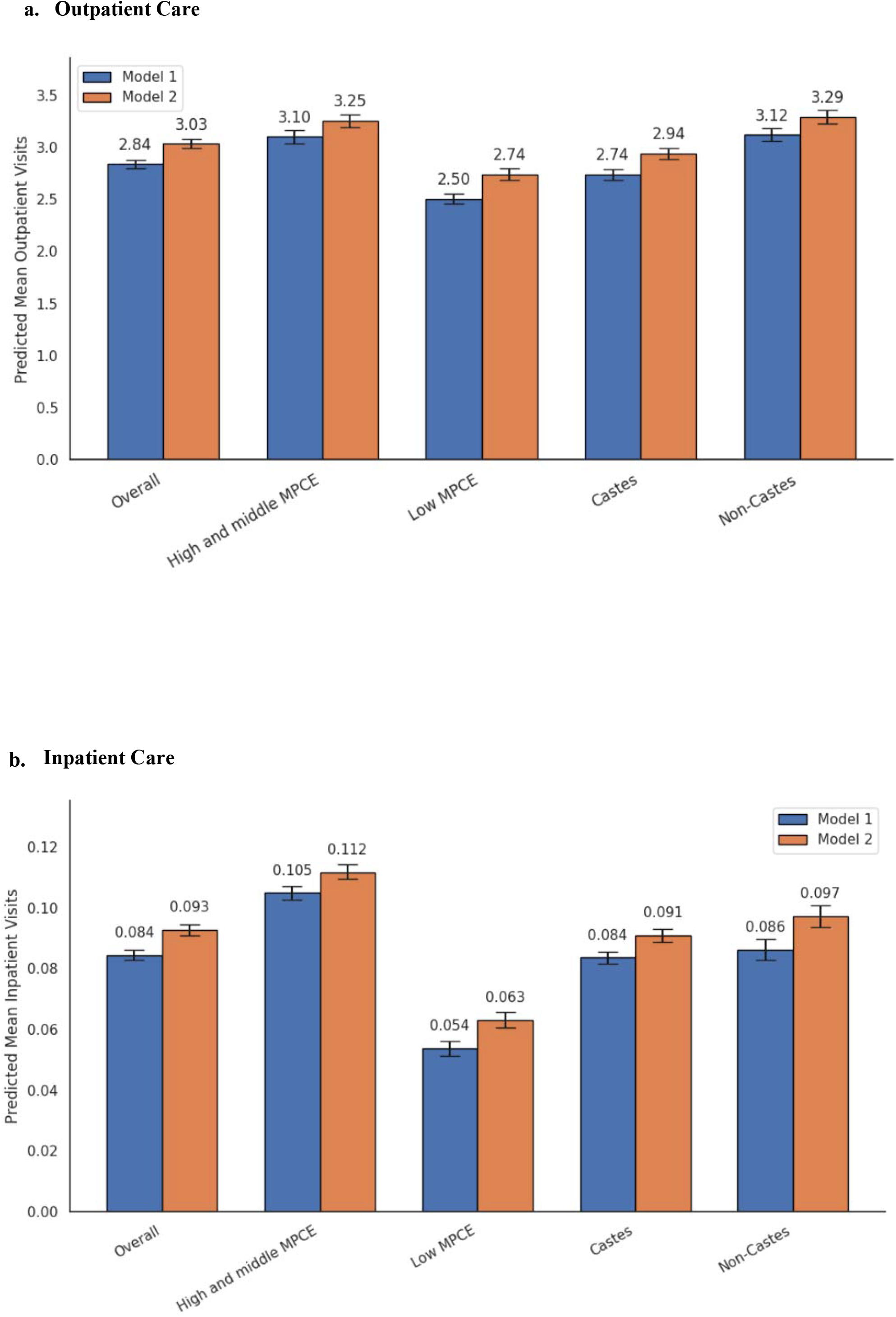
Expected Value Analysis (best performing weighted models)

**Appendix Figure 2.**
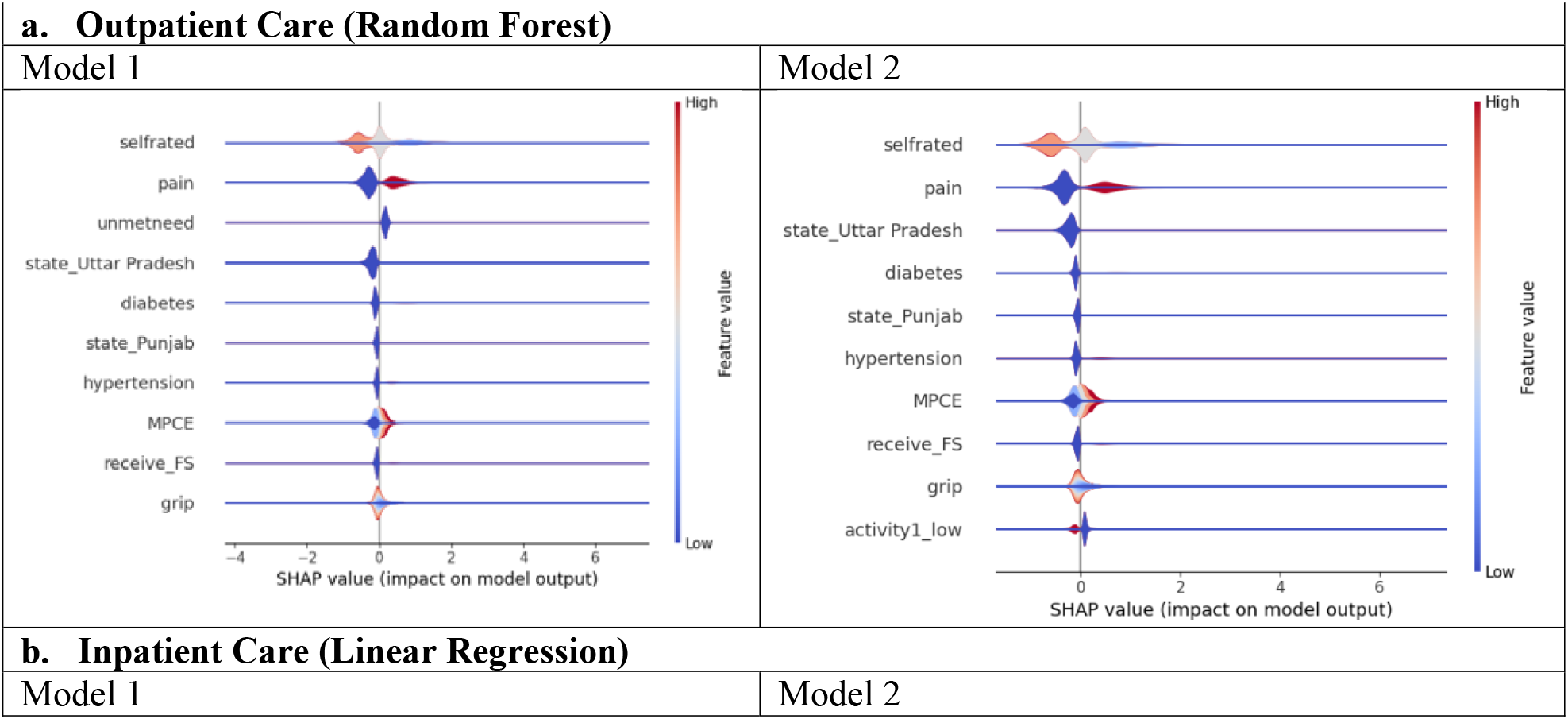
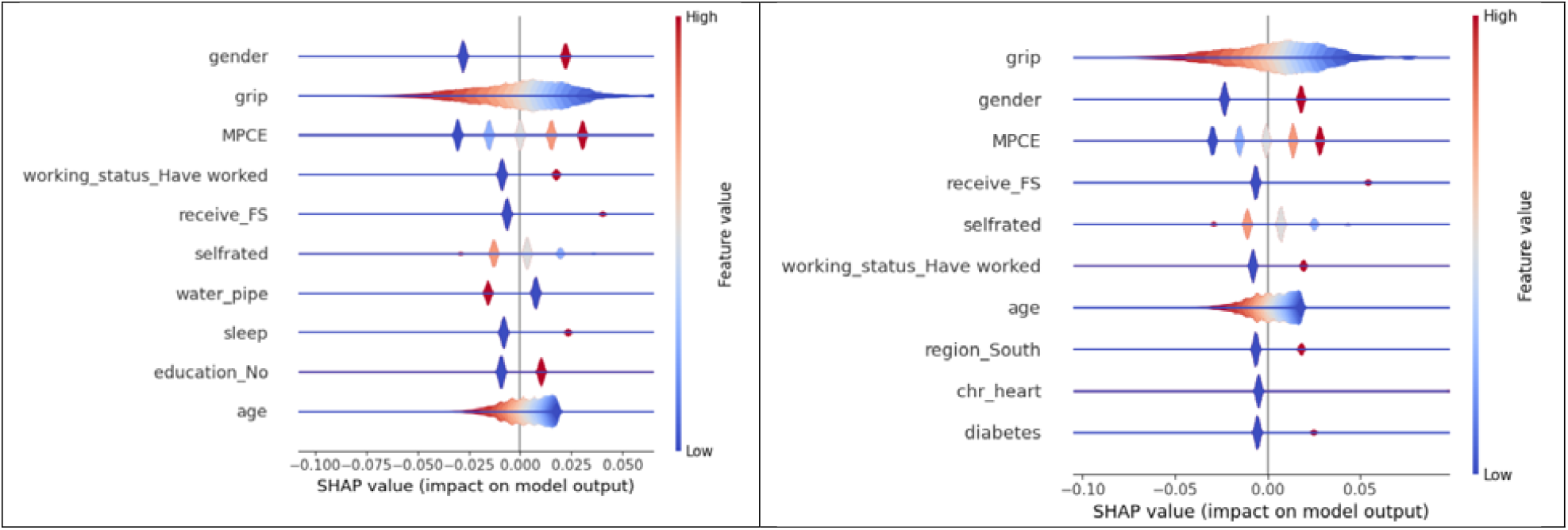
SHAP Value Analysis (best performing weighted models)

## REFERENCE

1. Mhasawade V, Zhao Y, Chunara R. Machine learning and algorithmic fairness in public and population health. Nat Mach Intell 2021; 3(8): 659–66.

2. Rajkomar A, Hardt M, Howell MD, Corrado G, Chin MH. Ensuring Fairness in Machine Learning to Advance Health Equity. Ann Intern Med 2018; 169(12): 866–72. Panch T, Mattie H, Atun R. Artificial intelligence and algorithmic bias: implications for health systems. J Glob Health. 2019 Dec;9(2):010318.

3. Panch T, Mattie H, Atun R. Artificial intelligence and algorithmic bias: implications for health systems. J Glob Health. 2019 Dec;9(2):010318.

4. Obermeyer Z, Powers B, Vogeli C, Mullainathan S. Dissecting racial bias in an algorithm used to manage the health of populations. Science 2019; 366(6464): 447-53.4.

5. Gianfrancesco MA, Tamang S, Yazdany J, Schmajuk G. Potential Biases in Machine Learning Algorithms Using Electronic Health Record Data. JAMA Intern Med 2018; 178(11): 1544–7.

6. Norori N, Hu Q, Aellen FM, Faraci FD, Tzovara A. Addressing bias in big data and AI for health care: A call for open science. Patterns (N Y) 2021; 2(10): 100347.

7. Ferryman K, Mackintosh M, Ghassemi M. Considering Biased Data as Informative Artifacts in AI-Assisted Health Care. N Engl J Med 2023; 389(9): 833–8.

8. Rajkomar A, Dean J, Kohane I. Machine Learning in Medicine. N Engl J Med 2019; 380(14): 1347–58.

9. Colacci M, Huang YQ, Postill G, et al. Sociodemographic bias in clinical machine learning models: a scoping review of algorithmic bias instances and mechanisms. J Clin Epidemiol 2025; 178: 111606.

10. World Health Organization. WHO CHOICE estimates of cost for inpatient and outpatient health service delivery. Economic Analysis and Evaluation Team, Department of Health Systems Governance and Financing. Geneva: World Health Organization; 2010. Available from: https://cdn.who.int/media/docs/default-source/health-economics/who-choice-estimates-of-cost-for-inpatient-and-outpatient-health-service-delivery.pdf

11. Stenberg K, Lauer JA, Gkountouras G, Fitzpatrick C, Stanciole A. Econometric estimation of WHO-CHOICE country-specific costs for inpatient and outpatient health service delivery. Cost Eff Resour Alloc 2018; 16: 11.

12. WHO. WHO-CHOICE estimates of cost for inpatient and outpatient health service delivery. 2021: 60.

13. Mehrabi N, Morstatter F, Saxena N, Lerman K, Galstyan A. A Survey on Bias and Fairness in Machine Learning. Acm Comput Surv 2021; 54(6).

14. Chen IY, Pierson E, Rose S, Joshi S, Ferryman K, Ghassemi M. Ethical machine learning in healthcare. Annual review of biomedical data science 2021; 4(1): 123–44.

15. Chen IY, Joshi S, Ghassemi M. Treating health disparities with artificial intelligence. Nature medicine 2020; 26(1): 16–7.

16. Adamson AS, Smith A. Machine Learning and Health Care Disparities in Dermatology. JAMA Dermatol 2018; 154(11): 1247–8.

17. Barocas S, Hardt M, Narayanan A. Fairness and machine learning: Limitations and opportunities: MIT press; 2023.

18. Zemel R, Wu Y, Swersky K, Pitassi T, Dwork C. Learning fair representations. International conference on machine learning; 2013: PMLR; 2013. p. 325–33.

19. Holstein K, Wortman Vaughan J, Daumé III H, Dudik M, Wallach H. Improving fairness in machine learning systems: What do industry practitioners need? Proceedings of the 2019 CHI conference on human factors in computing systems; 2019; 2019. p. 1–16.

20. Hardt M, Price E, Srebro N. Equality of opportunity in supervised learning. Advances in neural information processing systems 2016; 29.

21. Doshi-Velez F, Kim B. Towards a rigorous science of interpretable machine learning. arXiv preprint arXiv:170208608 2017.

22. Chen IY, Pierson E, Rose S, Joshi S, Ferryman K, Ghassemi M. Ethical Machine Learning in Healthcare. Annu Rev Biomed Data Sci 2021; 4: 123–44.

23. Panch T, Mattie H, Atun R. Artificial intelligence and algorithmic bias: implications for health systems. J Glob Health 2019; 9(2): 010318.

24. Veinot TC, Ancker JS, Bakken S. Health informatics and health equity: improving our reach and impact. J Am Med Inform Assoc 2019; 26(8-9): 689–95.

25. Reddy S, Fox J, Purohit MP. Artificial intelligence-enabled healthcare delivery. J R Soc Med 2019; 112(1): 22–8.

26. Miller DD, Brown EW. Artificial Intelligence in Medical Practice: The Question to the Answer? Am J Med 2018; 131(2): 129–33.

27. Organization WH. The implications of artificial intelligence and machine learning in health financing for achieving universal health coverage: findings from a rapid literature review. 2022.

28. Vayena E, Blasimme A, Cohen IG. Machine learning in medicine: Addressing ethical challenges. PLoS Med 2018; 15(11): e1002689.

